# Duration of BA.5 neutralization in sera and nasal swabs from SARS-CoV-2 vaccinated individuals, with or without Omicron breakthrough infection

**DOI:** 10.1101/2022.07.22.22277885

**Authors:** Delphine Planas, Isabelle Staropoli, Françoise Porot, Florence Guivel-Benhassine, Lynda Handala, Mathieu Prot, William-Henry Bolland, Julien Puech, Hélène Péré, David Veyer, Aymeric Sève, Etienne-Simon-Lorière, Timothée Bruel, Thierry Prazuck, Karl Stefic, Laurent Hocqueloux, Olivier Schwartz

## Abstract

Since early 2022, Omicron BA.1 has been eclipsed by BA.2, which was in turn outcompeted by BA.5, that displays enhanced antibody escape properties. Here, we evaluated the duration of the neutralizing antibody (Nab) response, up to 16 months after Pfizer BNT162b2 vaccination, in individuals with or without BA.1/BA.2 breakthrough infection. We measured neutralization of the ancestral D614G lineage, Delta and Omicron BA.1, BA.2, BA.5 variants in 291 sera and 35 nasal swabs from 27 individuals. Upon vaccination, serum Nab titers were reduced by 10-, 15-and 25-fold for BA.1, BA.2 and BA.5, respectively, compared with D614G. The duration of neutralization was markedly shortened, from an estimated period of 11.5 months post-boost with D614G to 5.5 months with BA.5. After breakthrough, we observed a sharp increase of Nabs against Omicron subvariants, followed by a plateau and a slow decline after 4-5 months. In nasal swabs, infection, but not vaccination, triggered a strong IgA response and a detectable Omicron neutralizing activity. Thus, BA.5 spread is partly due to abbreviated vaccine efficacy, particularly in individuals who were not infected with previous Omicron variants.

---

The SARS-CoV-2 Omicron BA.1 and BA.2 variants spread across the world and replaced the Delta variant in early 2022 ^1^. It is estimated that more than 50% of the population were infected by BA.1 or BA.2 by March 2022 ^2^ with a poor protection against infection conferred by vaccination ^3–5^. The incidence of breakthrough infections in vaccinated individuals has thus dramatically increased since Omicron emerged ^6^. BA.1 and BA.2 contain about 32 changes in the spike protein, promoting their high transmissibility and immune escape properties ^7–15^. The Omicron clade has rapidly evolved into sub-lineages, including BA.5, that outcompeted BA.1 and BA.2 ^16^. BA.5 spike shares multiple changes noted in BA.2 and bears a few additional modifications. BA.5 became predominant worldwide by mid-2022 and was responsible for a surge of contaminations in many countries ^16,17^. The neutralizing activity of sera from COVID-19 vaccine recipients is reduced against BA.5 by about 3-5 fold compared to BA.1 and BA.2 ^11,18–22^. Little is known about real-world vaccine efficacy against BA.5 infections, in individuals that were or were not previously infected with BA.1 or BA.2. Here, we assessed the durability and magnitude of neutralizing antibody (Nabs) responses against different Omicron variants, up to 16 months after Pfizer BNT162b2 vaccination. We also analyzed the evolution of Nabs in the sera and nasal swabs from vaccine recipients who experienced BA.1 or BA.2 breakthrough infections.

We longitudinally collected 291 sera and 35 nasal samples from a cohort of 27 health-care workers in Orleans, France. We previously studied the ability of some of these sera to neutralize Alpha, Beta, Delta and Omicron BA.1 variants ^10^. The characteristics of each participant are indicated in Supplemental table 1. 11 out of 27 individuals experienced a pauci-symptomatic breakthrough infection 60 to 178 days after the third injection. Screening by PCR or whole viral genome sequencing confirmed an Omicron breakthrough infection. At the time of infection (between December 2021 and mi-February 2022), BA.1 and BA.2 respectively represented 95% and 5% of the Omicron lineage in France ^23^. Each participant was sampled 3 to 21 times (mean=11) during the 16 months of survey. The days of vaccination, breakthrough infection and sampling are displayed Supplemental Figure 1.

We first measured serum Nab titers against the D614G reference virus, the Delta variant, and Omicron BA.1, BA.2 and BA.5 isolates in 22 vaccine recipients, 4-6 months after the booster doses. We calculated the 50% effective dilution (ED50) for each combination of serum and virus. As previously reported, Omicron subvariants displayed considerable immune escape properties, compared to D614G and Delta (Fig. 1a). Among Omicron subvariants, BA.5 neutralization was barely detectable, with a median ED50 of 60, reduced by a factor of 2.5 and 1.7 compared to BA.1 and BA.2 (ED50 of 148 and 100, respectively).

**Figure 1:**
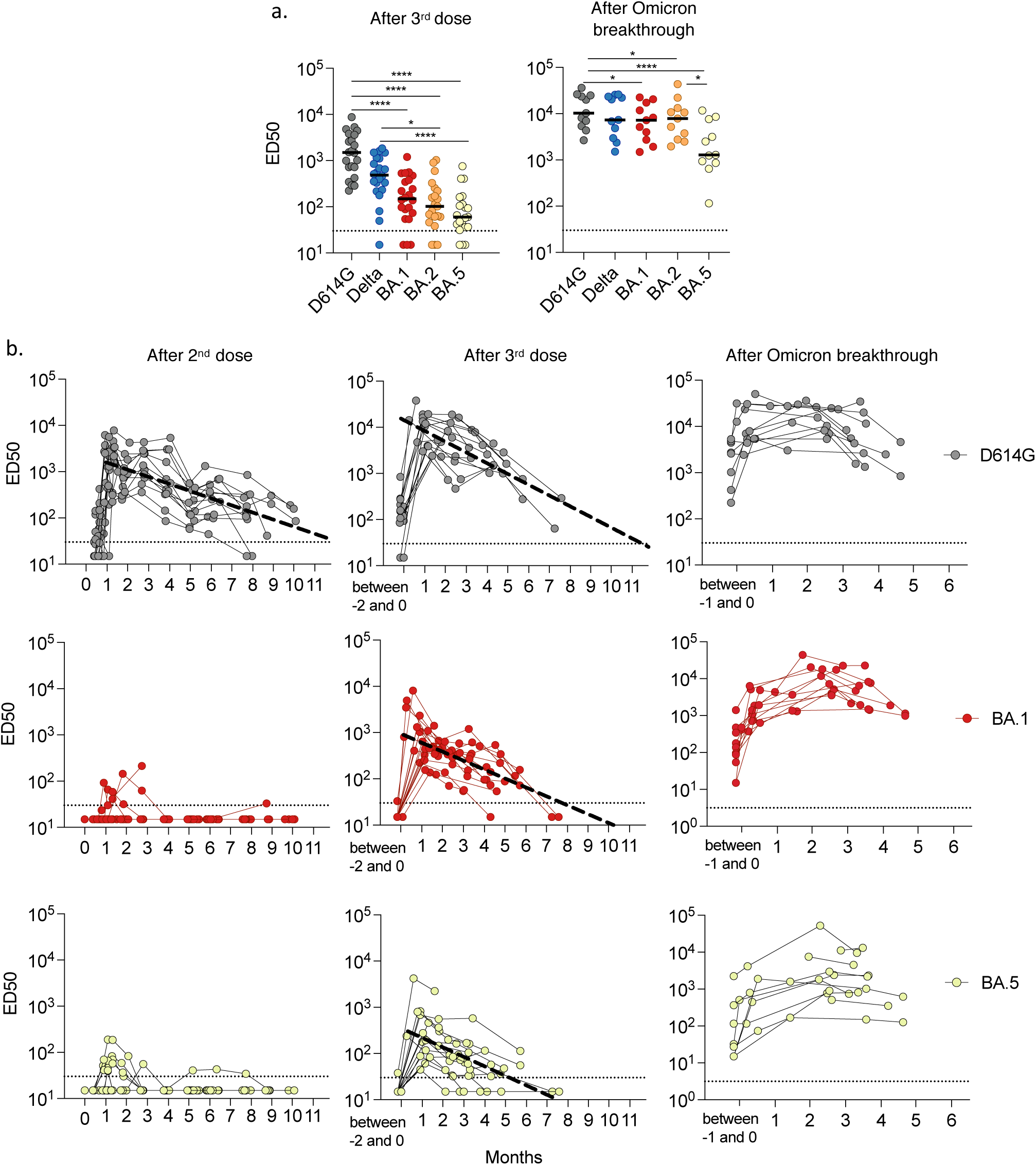
Magnitude, cross-reactivity and durability of antibodies in sera from Pfizer vaccinees, with or without breakthrough BA.1/BA.2 infection. **a**. Neutralizing antibody titers against D614G, Delta and Omicron BA.1, BA.2 and BA.5 were quantified in sera from triple vaccinated individuals (n= 22; median = 132 days post third dose) (left panel) and after Omicron BA.1 or BA.2 breakthrough infection (n=11; median = 80 days post infection). Two-sided Friedman test with Dunn’s test for multiple comparisons was performed between each viral strain at the different time points; *p < 0.05; **** p < 0.0001 **b**. Temporal evolution of Nab titers (ED50) against D614G (black), BA.1 (red) and BA.5 (yellow) in 27 vaccine recipients and 11 participants who had Omicron BA.1 or BA.2 breakthrough infection. The Nab titers were calculated at the indicated months after the second dose (left panels), third dose (middle panels) or breakthrough infection (right panels). The bold dotted line included in some panels represents a simple linear regression of Nab waning. In the other panels, the shape of the curves did not allow this analysis. Data are the mean from two independent experiments. The horizontal dotted line indicates the limit of detection (ED50 = 30).

One month after infection, the overall anti-SARS-CoV-2 IgG antibody levels increased by only 2.9 fold in the 11 individuals that experienced a breakthrough infection (Supplemental Figure 1). A strong augmentation of the cross-neutralization against Delta, BA.1 and BA.2 variants was observed, with ED50 ranging from 10^3^ to 10^4^ (Fig. 1a). The Nab titer was lower against BA.5 (ED50 of 10^3^). Therefore, post-vaccination infection by BA.1 or BA.2 led to an increase in Omicron-specific neutralizing antibody titers, which was less marked against BA.5.

We next longitudinally analyzed the evolution of serum cross neutralization in the 27 individuals, up to 16 months after initiation of the vaccination. We represented IgG levels and Nab titers at different time points after the second and third vaccine injection, as well as after Omicron BA.1 or BA.2 breakthrough infection. We performed modeling for the best fit curve of the data, using either continuous (simple linear regression), one-phase and two-phase decays. We compared the three decays using the extra sum-of-squares F Test, selecting the simpler model with p<0.05. The best fitting statistical model for the decline of anti-S IgG and Nab was the simple linear regression. As previously reported ^24^, a peak level of anti-Spike IgG was observed one month after the second dose, which subsequently declined over the next 10 months (Supplemental Fig. 2a). A booster dose induced higher IgG peak levels than the second dose. The regression model indicated that IgGs should become undetectable about 1.91 and 1.95 years after the second and third dose, respectively. The breakthrough infection increased IgG levels, without obvious decline up to 5 months, precluding calculation of a time to undetectability.

The evolution of the neutralization profile showed disparities between variants. After the second dose, we barely detected any neutralization against Omicron BA.1, BA.2 and BA.5, whereas D614G and to a lesser extent Delta were neutralized (Fig. 1a and supplemental Fig. 2). After boosting, the sera neutralized all variants, with differences not only in the peak level (Fig. 1a), but also in the duration of neutralization. The time-to-undetectability was shortened from 11.5 months for D614G, to 8 months for Delta and BA.1 and 5.5 months for BA.5 (Fig. 1a and supplemental Fig. 2). The kinetics of serum neutralization were different after breakthrough infection. With all variants, the peak was reached about two months after infection, followed by a slow decline not visible before 4-5 months.

**Figure 2:**
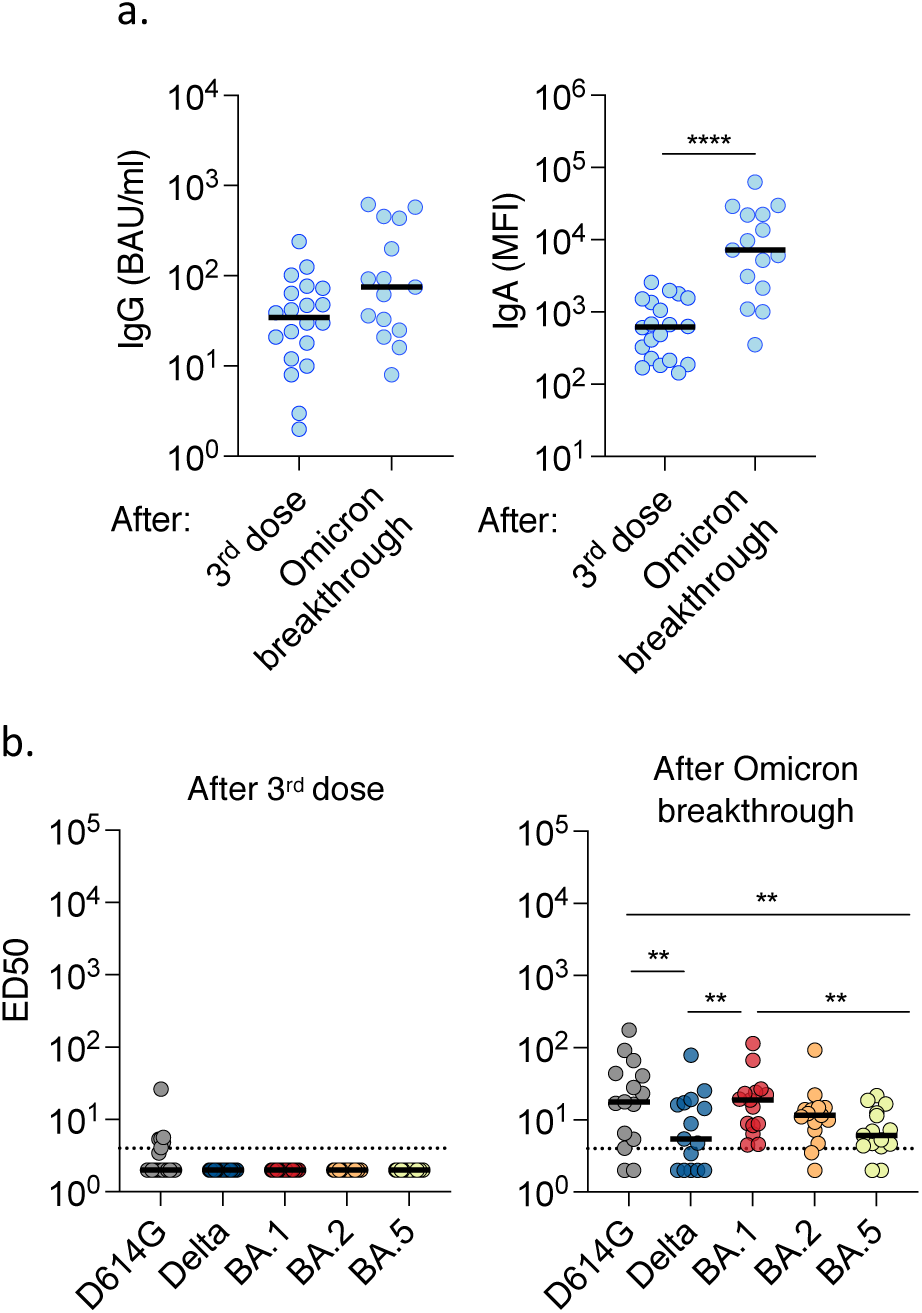
Induction of cross-neutralizing antibodies at the nasal mucosae upon Omicron BA.1 or BA.2 breakthrough infection. Nasal swabs were collected at 1 month post-third dose for n=25 participants (median = 38 days after the third dose) or 1 to 3 months after BA.1 or BA.2 breakthrough infection (n=7 participants sampled one to three times, representing a total of 15 samples). **a**. Levels of anti-spike IgGs (left panel) and IgAs (right panel) measured by flow cytometry with the S-Flow assay. For IgGs, results are presented in BAU/ml. The dotted lines indicate the limit of detection (3 BAU/ml). For IgAs, the mean fluorescence intensity (MFI) of binding is shown, since the lack of IgA reference samples precluded a calculation of BAU IgA/ml. Mann Whitney unpaired t-test was performed. ****p < 0.0001 **b**. The Nab titers against D614G, Delta and Omicron BA.1, BA.2 and BA.5 were measured in the same samples from triple vaccinated individuals (left panel) and after breakthrough infection (right panel). The dotted lines indicate the limit of detection (ED50 = 4). Two-sided Friedman test with Dunn’s test for multiple comparisons was performed between each viral strain at the different time points; **p < 0.01

To better visualize the influence of the lag between the booster and breakthrough infection on the evolution of cross-neutralization, we depicted the kinetics of Nab titers for 6 vaccine recipients with and 3 individuals without breakthrough infection (Supplemental Fig. 3). Breakthrough Omicron infections occurred 2 to 5 months post third dose and caused a consistent increase of anti-S IgGs and Nabs against the different variants. In non-infected individuals, Nabs declined progressively over the survey period.

Altogether, these results indicate a shorter neutralization efficacy of sera from triple vaccinated individuals against BA.5. A breakthrough Omicron infection triggered a longer lasting neutralizing response. There was no major impact of the timing of breakthrough infection relative to the vaccination on the extent of induction of cross-reactive Nabs.

We then asked whether vaccination and breakthrough infections may trigger different antibody responses at the mucosal surfaces. We measured the levels of anti-spike IgG and cross-neutralizing activity in 35 nasal swabs. 20 individuals were sampled one month after the third vaccine dose. Among them, seven experienced a breakthrough infection and were sampled one to three times, up to three months after infection, yielding a total of 15 samples. Levels of anti-SARS-CoV-2 IgG and IgA were relatively low (Fig. 2a), confirming that Pfizer vaccination does not induce a strong local immunity. The breakthrough infection triggered a moderate (2.2 fold) increase in IgGs, whereas IgAs were augmented by about 12 fold (Fig. 2a). Accordingly, in triple vaccinated individuals, we did not detect any neutralizing activity against Delta or Omicron variants, whereas only one individual poorly neutralized the D614G ancestral strain. In sharp contrast, 10 out of 15 nasal swabs collected after infection presented detectable neutralization against all variants. This neutralizing activity was higher against D614G, BA.1 and BA.2 than against Delta and BA.5. There was no obvious correlation between neutralization titers in the sera and nasal swabs (not shown). This is in agreement with a known compartmentalization of systemic and mucosal immune responses during acute SARS-CoV-2 infection ^25^. Therefore, Omicron breakthrough infection, but not vaccination, triggers a local Nab response at the viral entry site in the host. Our results help understand why a hybrid immunity is more effective than a vaccination to prevent infection by a novel SARS-CoV-2 variant.

The Omicron lineage has evolved towards enhanced transmissibility and immune evasion properties. BA.5 surged in many countries and displays a 3-5 fold lower sensitivity to Nabs generated by vaccination, relative to prior Omicron variants^18–21^. How this decrease impacts the duration of vaccine efficacy remains poorly characterized. Here, we longitudinally analyzed the levels of Nabs in a cohort of Pfizer vaccinated recipients. A booster dose generated a neutralizing response against the ancestral strain D614G that lasted about 11.5 months. The duration of neutralization was shortened to 5.5 months against BA.5, suggesting an abbreviated efficacy of current vaccines against this variant. In contrast, a natural Omicron infection in vaccinated individuals induced a longer lasting neutralizing response, with no visible decrease up to 4-5 months post infection. We also report that vaccination did not generate a detectable local neutralizing immunity at the nasal mucosae, whereas a breakthrough infection in vaccine recipients induced such a response. This suggests that the BA.5 wave preferentially occurred in “first-timer” individuals that were not previously infected with a prior Omicron variant, independently of their vaccination status.

Our results are in line with public health vaccine efficacy reports, analyzing tens of thousands of individuals in different countries ^26–29^. For instance, one study performed in England before the Omicron surge reported that two doses of Pfizer vaccine were associated with high short-term protection against SARS-CoV-2 infection^26^. This protection waned considerably after 6 months. Infection-acquired immunity boosted with vaccination remained high more than 1 year after infection ^27^. A study performed in Qatar reported that effectiveness of Pfizer vaccination against symptomatic BA.2 infection with two doses of Pfizer vaccine was negligible^28^, in agreement with our observation of a lack of detection Nabs against Omicron variants in sera from double vaccine recipients. The effectiveness of three doses of Pfizer, without or with previous infection was 52 % and 77%, respectively ^28^. Another report from USA indicated that vaccine effectiveness against COVID-19–associated hospitalization was higher during the BA.1 period than during the BA.2/BA.2.12.1 period ^29^.

Our study has limitations. The size of our cohort was relatively small, but the differences between variants were sufficiently marked to obtain statistical significance. We focused our work on Pfizer vaccine recipients and did not assess the neutralization conferred by a fourth dose. We analyzed the temporal evolution of the neutralizing response only up to 6 months post breakthrough infection. Future work will help understanding the efficacy of different vaccine regimen in various categories of individuals. It will be worth further examining the impact of a fourth vaccine dose on the extent and duration of the humoral immune response against BA.5 and other variants.

In summary, we show here that a longitudinal survey of the neutralizing humoral response in blood and nasal samples provides a reliable marker of the duration of effectiveness of vaccination, natural and hybrid immunity against current and forthcoming SARS-CoV-2 variants.

## Data Availability

All data produced in the present study are available upon reasonable request to the corresponding authors.

## Figure legends

**Supplemental Table 1.** Characteristics of the participants of the Pfizer vaccine recipient cohort.

| ID | Vaccines | Sex | Age | Date first-dose | Date second-dose | Date third-dose | Breakthrough | Variants |
| --- | --- | --- | --- | --- | --- | --- | --- | --- |
| #1 | Pfizer | F | 61-65 | Jan-21 | Jan-21 | Jul-21 | Dec-21 | BA.1 or BA.2 |
| #2 | Pfizer | M | 66-70 | Jan-21 | Jan-21 | Aug-21 | Dec-21 | BA.1 or BA.2 |
| #3 | Pfizer | M | 66-70 | Jan-21 | Jan-21 | Aug-21 | - | - |
| #4 | Pfizer | M | 56-60 | Jan-21 | Jan-21 | Jan-22 | - | - |
| #5 | Pfizer | M | 71-75 | Jan-21 | Mar-21 | Oct-21 | Jan-22 | BA.1 or BA.2 |
| #6 | Pfizer | M | 61-65 | Jan-21 | Jan-21 | Aug-21 | Jan-22 | BA.1 or BA.2 |
| #7 | Pfizer | M | 71-75 | Jan-21 | Jan-21 | - | - | - |
| #8 | Pfizer | F | unknown | Jan-21 | Jan-21 | Sep-21 | - | - |
| #9 | Pfizer | F | 51-55 | Jan-21 | Feb-21 | Nov-21 | - | - |
| #10 | Pfizer | F | 31-35 | Apr-21 | May-21 | Nov-21 | - | - |
| #11 | Pfizer | M | 61-65 | Jan-21 | Jan-21 | Sep-21 | Jan-22 | BA.1 or BA.2 |
| #12 | Pfizer | M | 41-45 | Jan-21 | Feb-21 | Nov-21 | Jan-22 | BA.1 |
| #13 | Pfizer | F | 36-40 | Jan-21 | Feb-21 | Oct-21 | Dec-21 | BA.1 |
| #14 | Pfizer | M | 51-55 | Jan-21 | Jan-21 | Aug-21 | - | - |
| #15 | Pfizer | F | 71-75 | Jan-21 | Jan-21 | - | - | - |
| #16 | Pfizer | F | 31-35 | Apr-21 | May-21 | Dec-21 | - | - |
| #17 | Pfizer | F | 56-60 | Jan-21 | Feb-21 | Nov-21 | - | - |
| #18 | Pfizer | M | 56-60 | Jan-21 | Feb-21 | Sep-21 | Feb-22 | BA.1 or BA.2 |
| #19 | Pfizer | F | 36-40 | Jan-21 | Jan-21 | Nov-21 | Jan-22 | BA.1 or BA.2 |
| #20 | Pfizer | M | 71-75 | Jan-21 | Jan-21 | Sep-21 | - | - |
| #21 | Pfizer | F | 56-60 | Jan-21 | Feb-21 | Nov-21 | - | - |
| #22 | Pfizer | F | 56-60 | Jan-21 | Feb-21 | Nov-21 | - | - |
| #23 | Pfizer | M | 56-60 | Jan-21 | Jan-21 | Aug-21 | Feb-22 | BA.1 or BA.2 |
| #24 | Pfizer | F | 91-96 | Feb-21 | Mar-21 | Sep-21 | - | - |
| #25 | Pfizer | M | 51-55 | Jan-21 | Jan-21 | Nov-21 | - | - |
| #26 | Pfizer | F | 71-75 | Jan-21 | Feb-21 | Nov-21 | - | - |
| #27 | Pfizer | M | 61-65 | Jan-21 | Jan-21 | Aug-21 | Jan-22 | BA.1 or BA.2 |

**Supplemental Figure 1:**
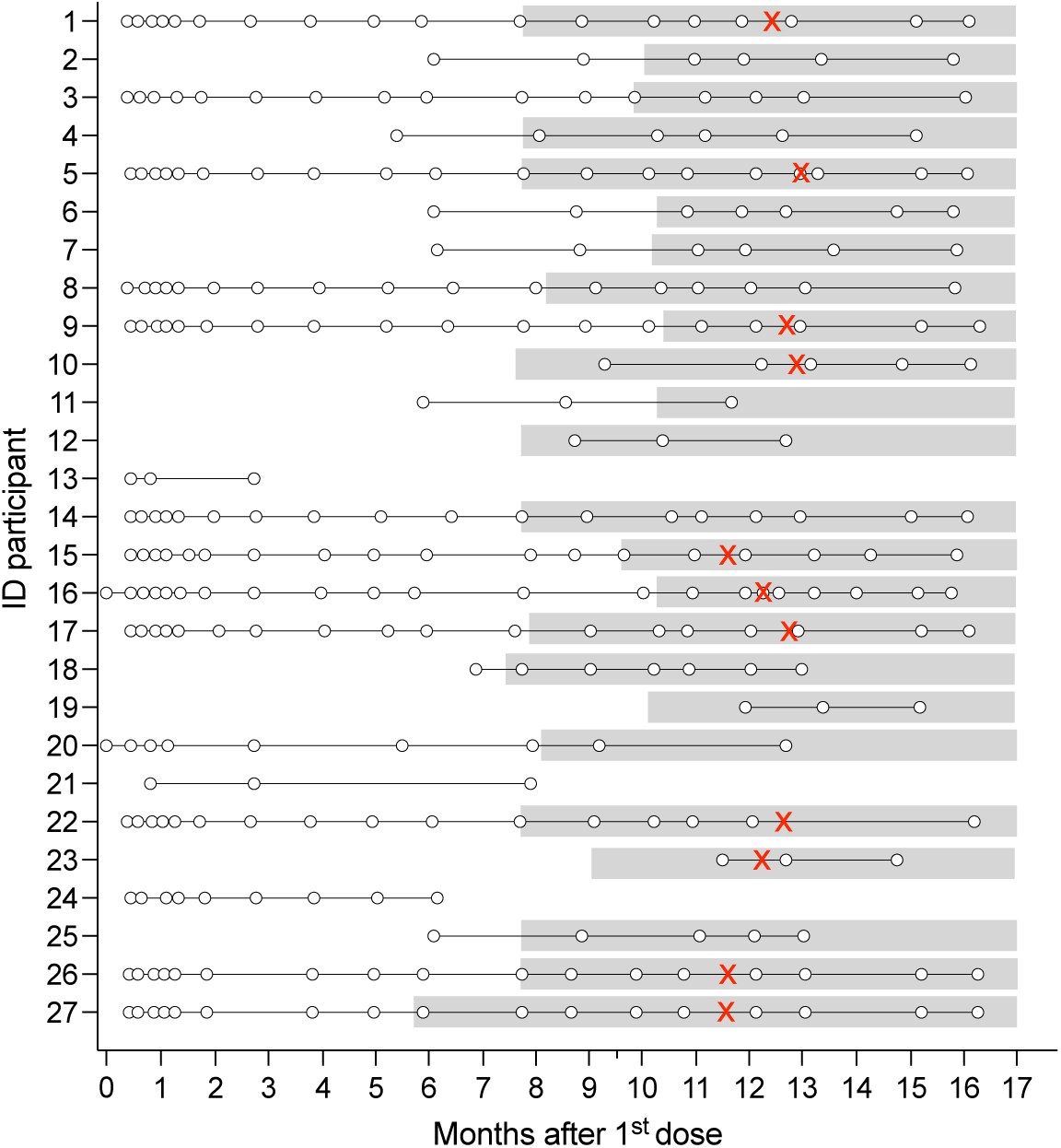
Chronology of vaccination and breakthrough infection. The 27 participants received 2 or 3 doses of Pfizer BNT162b2 vaccines. Sera were collected at the indicated time points after the first dose (white circles). The gray background indicates the period of time following the third dose. (x-axis is the timeline post first injection. The red crosses represent the occurrence of BA.1 or BA.2 breakthrough infection in 11 participants.

**Supplemental Figure 2:**
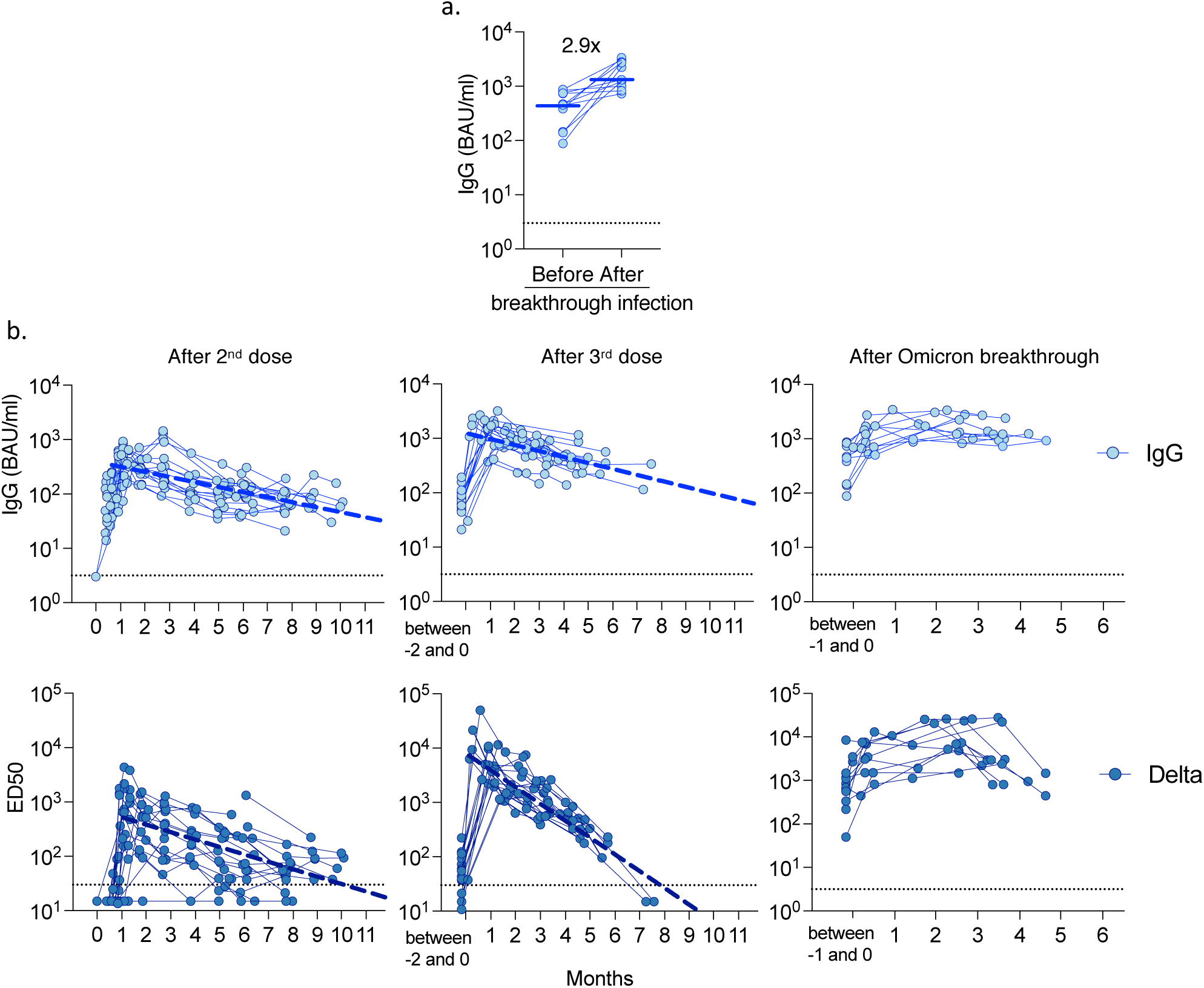
Magnitude, cross-reactivity and durability of antibodies in sera from Pfizer vaccinees, with or without breakthrough BA.1/BA.2 infection. **a**. Levels of anti-spike IgGs measured by flow cytometry with the S-Flow assay before (median = 20 days) and after (median = 80 days) in 11 individuals Omicron breakthrough. Results are presented in BAU/ml. The dotted lines indicate the limit of detection (3 BAU/ml). **b**. Temporal evolution of IgG levels (upper panels) and Nab titers (ED50) against Delta in 27 vaccine recipients and 11 participants who had Omicron BA.1 or BA.2 breakthrough infection. The IgG levels and Nab titers were calculated at the indicated months after the second dose (left panels), third dose (middle panels) or breakthrough infection (right panels). The bold dotted line included in some panels represents a simple linear regression of Nab waning. In the other panels, the shape of the curves did not allow this analysis. Data are the mean from two independent experiments. The horizontal dotted line indicates the limit of detection (ED50 = 30).

**Supplemental Figure 3:**
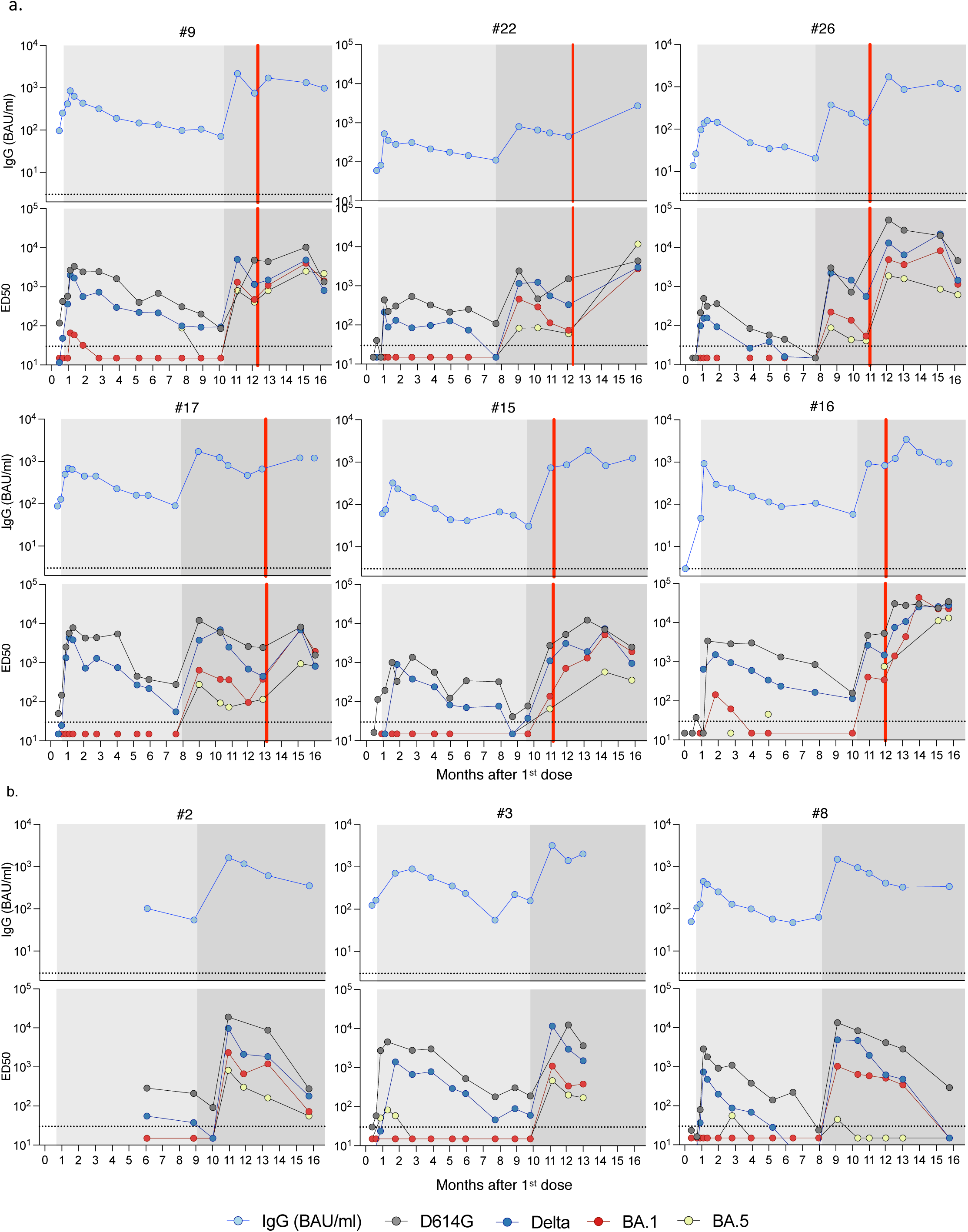
Temporal evolution of anti-Spike IgG and Nab titers after vaccination in nine individuals from the cohort. We selected individuals with a large number of available longitudinal samples. Six individuals with Omicron breakthrough (a) infection and three individuals without Omicron breakthrough infection (b) are depicted. For each individual, the hashtag number corresponds to the ranking in supplemental Table 1. The upper panels represent the evolution of IgG levels, and the lower panels the neutralization profile against the indicated variants. The white, light grey and dark grey backgrounds indicate the period of time with one, two or three 3 doses of vaccine. The red line corresponds to the last sample collected before breakthrough infection. Data are the mean from two independent experiments. The dotted lines indicate the limit of detection (BAU/ml = 3; ED50 = 30).

**Supplemental Figure 4:**
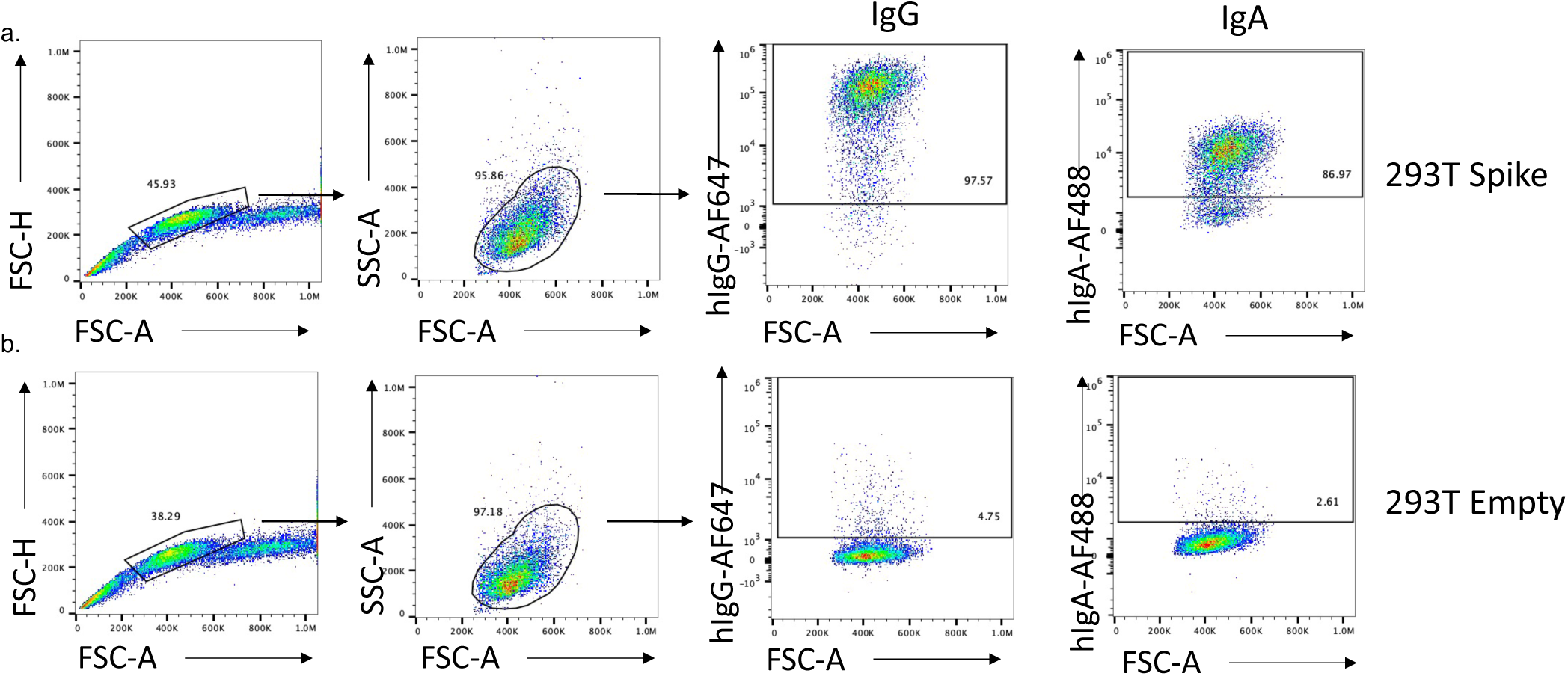
Gating strategy of the S-Flow assay. 293T cells stably expressing the Wuhan Spike were incubated with sera from vaccinated individua (dilution 1:300) and stained with a mix of anti-human IgG (AlexaLFluor 647) plus anti-human IgA (AlexaFluor 488) and analyzed by flow-cytometry. (a) One representative example of the gating strategy for anti-IgG and IgA is shown. (b) Gates are set on cells transfected with a control plasmid not encoding a spike (293T empty). An example of the signal obtained by a reactive serum on spike expressing cells is shown.

## Methods

No statistical methods were used to predetermine sample size. The experiments were not randomized, and the investigators were not blinded to allocation during experiments and outcome assessment. Our research complies with all relevant ethical regulation.

## Cohort

We conducted a prospective, monocentric, longitudinal, interventional cohort clinical study (ABCOVID) since 27 August 2020 with the objective to study the kinetics of COVID-19 antibodies in patients with confirmed SARS-CoV-2 infection (NCT04750720). A sub-study aimed to describe the kinetic of neutralizing antibodies (in blood and nasal mucosae) after vaccination. The cohort was previously described^10,30,31^. The participants, that were not previously infected at the time of inclusion, received two doses of Pfizer BNT162b2 vaccine within an interval of 21-28 days and a booster dose 154 to 361 days later. This study was approved by the Ile-de-France IV ethical committee. At enrollment, written informed consent was collected and participants completed a questionnaire covering sociodemographic characteristics, virological findings (SARS-CoV-2 RT–qPCR results, date of testing) and data related to SARS-CoV-2 vaccination (brand product, date of first, second and third vaccination). Blood and nasal swabs were collected at several time points. Nasal swabs were collected in the two cavities and preserved in 1 or 3 ml of M4RT transport buffer (Standard Sigma Swabs MW910S, Sigma VCM). Study participants did not receive any compensation.

## S-Flow Assay

The S-Flow assay is based on the recognition of the SARS-CoV-2 spike protein expressed on the surface of 293T cells. It was used to quantify SARS-CoV-2-specific IgG and IgA subtypes in sera, nasopharyngeal swab supernatants and saliva as previously described ^32,33^. Briefly, 293T cells were obtained from ATCC (ATCC Cat# CRL-3216, RRID:CVCL_0063) and tested negative for mycoplasma. 293T cells stably expressing Spike (293T S) or control (293T Empty) cells were transferred into U-bottom 96-well plates (105 cells/well). Cells were incubated at 4°C for 30 min with serum (1:300 dilution) or nasal swabs (1:50) in PBS containing 0.5% BSA and 2 mM EDTA. Then, cells were washed with PBS, and stained using anti-IgG AlexaFluor647 (Jackson ImmunoResearch cat# 109-605-170) and Anti-IgA AlexaFluor488 (Jackson ImmunoResearch cat# 109-545-011). Cells were washed with PBS and fixed for 10 min with 4% PFA. Data were acquired on an Attune Nxt instrument (Life Technologies). Stainings were also performed on control (293T Empty) cells. Results were analyzed with FlowJo 10.7.1 (Becton Dickinson). The gating strategy for IgGs and IgAs is shown in Supplemental Figure 4. The specific binding was calculated as follows: 100 x (% binding 293T Spike - % binding 293T Empty)/ (100 - % binding 293T Empty). The assay was standardized with WHO international reference sera (20/136 and 20/130) and cross-validated with two commercially available ELISA (Abbott and Beckmann) using a Passing-Bablok linear regression model to allow calculation of BAU/mL^34^.

## S-Fuse neutralization assay

U2OS-ACE2 GFP1-10 or GFP 11 cells, also termed S-Fuse cells, become GFP+ when they are productively infected by SARS-CoV-2 ^31,35^. Cells were tested negative for mycoplasma. Cells were mixed (ratio 1:1) and plated at 8×10^3^ per well in a μClear 96-well plate (Greiner Bio-One). The indicated SARS-CoV-2 strains were incubated with serially diluted sera (first dilution 1:30) or nasal swabs (first dilution 1:4) for 15 minutes at room temperature and added to S-Fuse cells. The sera were heat-inactivated for 30 min at 56°C before use. 18 hours later, cells were fixed with 2% PFA, washed and stained with Hoechst (dilution 1:1,000, Invitrogen). Images were acquired with an Opera Phenix high content confocal microscope (PerkinElmer). The GFP area and the number of nuclei were quantified using the Harmony software (PerkinElmer). The percentage of neutralization was calculated using the number of syncytia as value with the following formula: 100 x (1 – (value with serum – value in “non-infected”)/(value in “no serum” – value in “non-infected”)). Neutralizing activity of each serum was expressed as the half maximal effective dilution (ED50). ED50 values (dilution values for sera and nasal swabs) were calculated with a reconstructed curve using the percentage of the neutralization at the different concentrations.

## Virus strains

The reference D614G strain (hCoV-19/France/GE1973/2020) was supplied by the National Reference Centre for Respiratory Viruses hosted by Institut Pasteur (Paris, France) and headed by Pr. S. van der Werf ^31^. This viral strain was supplied through the European Virus Archive goes Global (Evag) platform, a project that has received funding from the European Union’s Horizon 2020 research and innovation program under grant agreement n° 653316. Delta was isolated from a nasopharyngeal swab of a hospitalized patient returning from India ^30^. The swab was provided and sequenced by the laboratory of Virology of Hopital Européen Georges Pompidou (Assistance Publique – Hopitaux de Paris). The Omicron BA.1 and BA.2 strains were supplied and sequenced by the NRC UZ/KU Leuven (Leuven, Belgium)^36^. The Omicron BA.5 was isolated from a nasopharyngeal swab of woman hospitalized for rheumatological pain. The swab was sequenced by the National Reference Center for HIV-Associated laboratory of Tours, France. All patients provided informed consent for the use of the biological materials.

The variant strains were isolated from nasal swabs using Vero E6 cells and amplified by one or two passages. Viruses were sequenced directly on nasal swabs, and after one or two passages on Vero cells. Sequences of the swabs and amplified viruses were similar. Sequences were deposited on GISAID immediately after their generation, with the following IDs: D614G: EPI_ISL_41463; Delta ID: EPI_ISL_2029113; Omicron BA.1 ID: EPI_ISL_6794907; Omicron BA.2 GISAID ID: EPI_ISL_10654979; Omicron BA.5 ID: EPI_ISL_13660702. Titration of viral stocks was performed on Vero E6, with a limiting dilution technique allowing a calculation of TCID50, or on S-Fuse cells.

## Statistical analysis

Flow cytometry data were analyzed with FlowJo v10 software (TriStar). Calculations were performed using Excel 365 (Microsoft). Figures were drawn on Prism 9 (GraphPad Software). Statistical analysis was conducted using GraphPad Prism 9. Statistical significance between different groups was calculated using the tests indicated in each figure.

## Author contributions

Experimental strategy design, experiments: D.P., I.S., F.P., F.G.-B., W.-H.B., T.B., M.P., E.S.-L. and O.S.

Vital materials: L.H., J.P., H.P., D.V., A.S., T.P., K.S., L.H.

Manuscript writing: D.P. and O.S.

Manuscript editing D.P., W.-H. B., E. S-L., T.B. and O.S.

## Acknowledgments

We thank A. Fontanet for critical reading of the manuscript, the patients who participated to this study, members of the Virus and Immunity Unit and other teams for discussion and help, N. Aulner and the staff at the UtechS Photonic BioImaging (UPBI) core facility (Institut Pasteur), a member of the France BioImaging network, for image acquisition and analysis, F. Peira, V. Legros and L. Courtellemont for their help with the cohorts.

